# Molecular and phylogenetic insights into the novel *Brugia* sp. in Sri Lanka with new evidence for zoonotic transmission

**DOI:** 10.64898/2026.07.20.26358473

**Authors:** Sachini U. Nimalrathna, Hiruni Harischandra, Michael J. Kimber, Nilmini Chandrasena, Nilanthi de Silva, Chandana H. Mallawarachchi, B. G. D. Nissanka K. de Silva

## Abstract

The World Health Organization (WHO) validated Sri Lanka had eliminated lymphatic filariasis as a public health problem in 2016, the second country in Southeast Asia to attain this status. However, post-validation surveillance has identified sporadic cases of brugian filariasis. The reemergence of *Brugia malayi* infections in Sri Lanka warrants urgent investigations. Recent studies have shown that the parasite responsible for the reemergence is a novel zoonotic *Brugia* sp. maintained among dogs that is closely related but distinct to the human-infecting *B. malayi* species. The current study employed morphological and morphometric assessments, revealing that this novel zoonotic *Brugia* sp. is within the *B. malayi* morphological range. Molecular characterization of three genomic regions, the nuclear genomic region *SLXI*, the non-coding region HhaI, and the mitochondrial genomic region *COXI* confirmed it as a genetic variant more closely related to *B. malayi* than to *B. pahangi*. Phylogenetic analysis further indicated it as a distinct genomic variant, closely related to a *B. malayi*-like parasite reported from India. Notably, that same parasite was identified in infected humans, animals, and potential vector mosquitoes. This, together with the detection of both human and animal blood within the same brugian infective mosquitoes, and delineating the canine origin of the parasites in human infections, provides compelling evidence supporting zoonotic transmission of this parasite. To our knowledge, this is the first report demonstrating the presence of the same brugian parasite in humans, domestic animals, and potentially infective mosquitoes in Sri Lanka, supported by multi-genomic evidence. The recent identification of multiple potential mosquito vector species suggests that this parasite may have undergone adaptive changes, facilitating its ability to overcome the species barrier. These findings substantiate the long-held hypothesis of zoonotic transmission of the reemerged brugian parasite, highlighting significant implications for ongoing surveillance and control strategies.

**Author Summary:** Brugian filariasis was eradicated from Sri Lanka in 1969 but has reemerged. The parasite driving this reemergence remains unclear, but recently, a novel, potentially zoonotic *Brugia* sp. has been identified. The current study morphologically and molecularly characterized this human brugian parasite. Morphologically, the parasite was identified as *Brugia* sp., with body measurements corresponding to the size range of previously reported *Brugia malayi*. Molecular analysis revealed higher similarity to *B. malayi* than to *B. pahangi*. Phylogenetic analysis revealed close similarity to another novel *B. malayi*-like parasite reported from India. Analysis of brugian parasites across humans, animals, and mosquitoes identified the same brugian parasite within humans, cats, dogs, and mosquitoes. Additionally, blood meal analysis of potentially infective mosquitoes further supported zoonotic transmission of the disease. These potential vector mosquitoes breed in different sites, and their biting and resting behaviors also vary. As a result, conventional vector control methods are often ineffective. In addition, the presence of animal reservoir species (cats and dogs) has contributed to active transmission of the parasite among suburban and rural human populations. Therefore, reemergence of a genetic variant brugian parasite and zoonotic transmission is a concern for health authorities in terms of surveillance and planning new control strategies.

## Introduction

Lymphatic filariasis (LF) is a vector-borne, Neglected Tropical Disease (NTD) caused by parasitic filarial nematodes, affecting 657 million people in 39 countries, including Sri Lanka [1]. The most characteristic clinical manifestation of LF is elephantiasis, marked by severe swelling of the extremities resulting from parasite-induced obstruction of the lymphatic system. This condition makes LF a leading cause of permanent physical disfigurement and one of the major causes of long-term disability worldwide [1,2]. Severe morbidity from this disease imposes a socioeconomic burden and hinders community development in developing countries [3–7], resulting in an annual economic burden of over US $5.8 billion for treatment, healthcare costs, and potential income loss [8]. In Sri Lanka, 52% (11,200,000) of the population lives in the former LF endemic region and may be at risk of infection [9–13].

LF can be subdivided by the species of filarial nematode causing the infection. Bancroftian filariasis is caused by *Wuchereria bancrofti,* and brugian filariasis in humans is mainly caused by *Brugia malayi,* and *B. timori,* and in animals by *B. pahangi*. Bancroftian filariasis is prevalent in tropical Africa, Southeast Asia, Papua New Guinea, the Philippines, and Thailand, while brugian filariasis is localized to South and Southeast Asia, including Sri Lanka [14]. Historically, Sri Lanka experienced the coexistence of both brugian and bancroftian filariasis, often described as “rural” and “urban” types, respectively. Brugian filariasis was eliminated from Sri Lanka in the late 1960s through vector control activities targeting the breeding sites of *Mansonia* spp. mosquitoes (clearing aquatic vegetation). Aligning with the WHO’s Global Programme to Eliminate Lymphatic Filariasis (GPELF), Sri Lanka commenced the National Programme to Eliminate LF in 2002 [13].

As a result of the annual Mass Drug Administration (MDA) programme coordinated by GPELF, Sri Lanka received its LF elimination status in 2016, a momentous achievement for the nation [15]. However, post-MDA surveillance has revealed sporadic cases of human brugian filariasis in all LF-endemic provinces in Sri Lanka [16,17]. Additionally, several studies have reported a surge in brugian filariasis among canine and feline populations in Sri Lanka over the past fifteen years, suggesting that dogs and cats are reservoir hosts for brugian filariasis in the country [17–19]. Filarial parasites in both dog and cat populations have been intensively studied in other South and Southeast Asian countries, and both have been implicated as reservoir hosts for LF due to the high prevalence of microfilaraemia [20,21]. This raises concerns about whether the recent increase in microfilaraemic rates among animals reflects the presence of a parasite with zoonotic potential. Supporting this, a recent investigation revealed that brugian parasites in a human case from Gampaha, Western Province, Sri Lanka, exhibit sub-periodicity, a feature of zoonotic brugian filariasis [16]. Studies have also found that the brugian parasite responsible is morphometrically and phylogenetically distinct from the previously characterized nocturnally periodic brugian parasite in Sri Lanka [16,17,22,23]. Additionally, studies report that the prevailing brugian parasite is distinct from the *B. malayi* parasite and shows higher sequence homology to *B. pahangi* than to *B. malayi,* with corresponding phylogenetic evidence [17]. Mallawarachchi *et al.* have reported the low sensitivity of the *Brugia* Rapid Test (BRT), recommended by the WHO as a rapid screening tool for brugian filariasis, in detecting *Brugia* spp. microfilaraemic human cases, based on a study conducted in the Gampaha district, Sri Lanka [16]. Recently, a study has reported several additional potential vector mosquito species for brugian filariasis in Sri Lanka [24]. Collectively, these findings suggest the emergence of a new species or a genetic modification of the existing *B. malayi*, warranting further investigation.

The present study aimed to investigate the prevailing brugian parasite infecting humans in Sri Lanka. Morphological, morphometric, molecular, and phylogenetic approaches were employed for parasite characterization. Three genomic regions of the parasite were compared with existing brugian parasite data and with lab-raised brugian parasites to assess genetic variations through molecular and phylogenetic analyses. Furthermore, the zoonotic potential of the parasite was investigated by evaluating its ability to cross species barriers and establish infections in feline and canine populations. In addition, potential vector mosquitoes were screened for brugian parasites, and their blood meals were analyzed to investigate the zoonotic transmission of the disease. Finally, the likely origin of the parasite was inferred through comparative gene analysis in conjunction with existing literature.

## Methods

### Ethics approval

Ethical clearance for the human and animal subjects of the study was obtained from the Ethics Review Committees (ERC) of the Faculty of Medicine, University of Sri Jayewardenepura (03/20) and the Institute of Biology, Sri Lanka (IOBSL) (ERC IOBSL 206.02.2020), respectively. Approval to conduct the study in each province was obtained from the respective Regional Director of Health Services (RDHS) and Divisional Veterinary Officers.

### Collection of human blood samples

Venous blood samples (n=14) were obtained from the Anti-filariasis Campaign (AFC) Headquarters, Narahenpita, Sri Lanka, which were collected from humans identified as brugian filariasis infected by Giemsa-stained thick blood smears (TBSs) observation. The blood samples were collected between 22:00 and 24:00 Sri Lankan standard time during routine surveillance activities in five filariasis-endemic areas. Laboratory-reared and maintained *B. malayi* and *B. pahangi* samples were obtained from the Filariasis Research Reagent Resource Center (FR3) at the University of Georgia, United States of America, and were used as reference samples for this study.

### Morphological identification of the *Brugia* sp. in humans

TBSs were prepared according to standard protocols using 60 μl of *Brugia* sp. infected blood [25], Giemsa-stained, and observed by light microscope (Biomed, LB-202). Body measurements (mean total body length, mean length of the cephalic space, mean width of the cephalic space, and the cephalic space width-to-length ratio) were measured using a stereo microscope (Biomed, LB-202) and the Image Analysis software version 5.1.

Statistical analysis of morphometric measurements was performed using Minitab 17 software (Minitab, State College, PA, USA). One-way *ANOVA* was conducted to evaluate variability in each morphometric measurement across the samples. Significant variables from all measured variables were selected for principal component analysis (PCA). The selection of principal components in the PCA was justified based on the eigenvalues and the cumulative correlation of the total components.

### Molecular and phylogenetic analysis of the *Brugia* sp. in humans

DNA was extracted from *Brugia* sp. infected blood using the Blood and Tissue DNA Extraction Kit (Qiagen, Germany) according to the manufacturer’s guidelines with some modifications. DNA was eluted in nuclease-free water in two subsequent steps, adding 20 μL and 15 μL and centrifuging at 8,000 rpm for 1 min after each addition. DNA concentration was determined using a NanoDrop Spectrophotometer (Thermo Fisher Scientific, Waltham, MA, USA).

Three regions of the brugian parasites in infected human blood samples, namely the Cytochrome c Oxidase subunit I (*COXI*) region, HhaI repeat region, and Trans-Spliced Leader Exon I (*SLXI*) repeat regions, were selected for sequence comparison. The 517 bp and 633 bp *COXI* regions of *B. malayi* and *B. pahangi* were amplified, respectively, using forward primer 5′-GGACCAGGAAGTAGTT-3′, reverse primer 5′-TATACATATGGTGACCTC-3′, and forward primer 5′-TATTGCCTGTTATGC-3′, reverse primer 5′-TGTATATGTGATGAC-3′ as previously described [26]. Cycling conditions were 95 °C for 10 mins followed by 30 cycles of 1 min at 94 °C, 1 min at 54 °C and 2 mins at 72 °C followed by 10 mins at 72 °C. The 322 bp *Brugia* species-specific HhaI repeat region was PCR amplified as previously described [27] using 5’-GCGCATAAATTCATCAGC-3’ (Forward) and 5’-GCGCAAAACTTAATTACAAAAGC-3’ (Reverse) primers. Cycling conditions were 94 °C for 5 mins followed by 35 cycles each of 1 min at 94 °C, 1 min at 59 °C and 1 min at 72 °C followed by 10 mins at 72 °C. The 294 bp *SLXI* region of *Brugia* sp. was amplified using the region-specific forward primer 5’-GTCTACGACCATACCACGTTGA-3’and the reverse primer 5’-GAAACATTCAATTACCTCAAAC–3’as previously described [20]. Cycling conditions were 95 °C for 4 mins followed by 30 cycles of 30 s at 95 °C, 30 s at 59 °C and 45 s at 72 °C and finally 10 mins at 72 °C.

Each region was sequenced using Sanger sequencing [28] at either the Iowa State DNA sequencing facility (Iowa State University, Ames, IA, USA) or Macrogen (Seoul, Republic of Korea). PCR products with DNA concentrations insufficient for sequencing were sub-cloned into the pGEM-T Easy vector (Promega, Madison, WI, USA), transformed into JM109 competent cells (Promega, Madison, WI, USA), plasmids were extracted with PureYield Plasmid Miniprep kit (Promega, Madison, WI, USA), and sequenced. The sequences were initially analyzed using BLAST (Basic Local Alignment and Searching Tool), and base-by-base analysis was performed using Multiple Sequence Alignment (MSA) with FR3 sequences of *B. malayi* and *B. pahangi* using CLC Main Workbench 23.0.1 (Qiagen, Germany). Bayesian Phylogeny of parasite lineages was constructed using MrBayes version 3.2 and BEAST version 1.10.4, and the phylogenetic tree was visualized using FigTree version 1.4.4. Kimura 2-parameter (K2P) genetic divergence analysis was conducted with the *COXΙ* genomic sequences of the parasites.

### Collection of mosquitoes and animal blood samples

High-risk districts were identified based on the number of human brugian filariasis cases reported to the AFC. A 500 m radius zone around the most recent human brugian filariasis case in each district was demarcated as the study site (Fig 1). The selected study sites were Puruduwella-Puttalam District (S_1_) (7.4898707, 79.859083), Maggona-Kalutara District (S_2_) (6.51012, 79.992529), Wattala-Gampaha District (S_3_) (6.99634, 79.89580), Induruwa-Galle District (S_4_) (6.3771134, 80.0171609), Boralesgamuwa-Colombo District (S_5_) (6.86579, 79.87821) and Mahawewa-Puttalam District (S_6_) (7.465026, 79.8697312). The buffer zone was chosen to encompass the mean flight range of *Mansonia* spp. mosquitoes (350 m), the known endemic vector for *B. malayi*.

**Fig 1.**
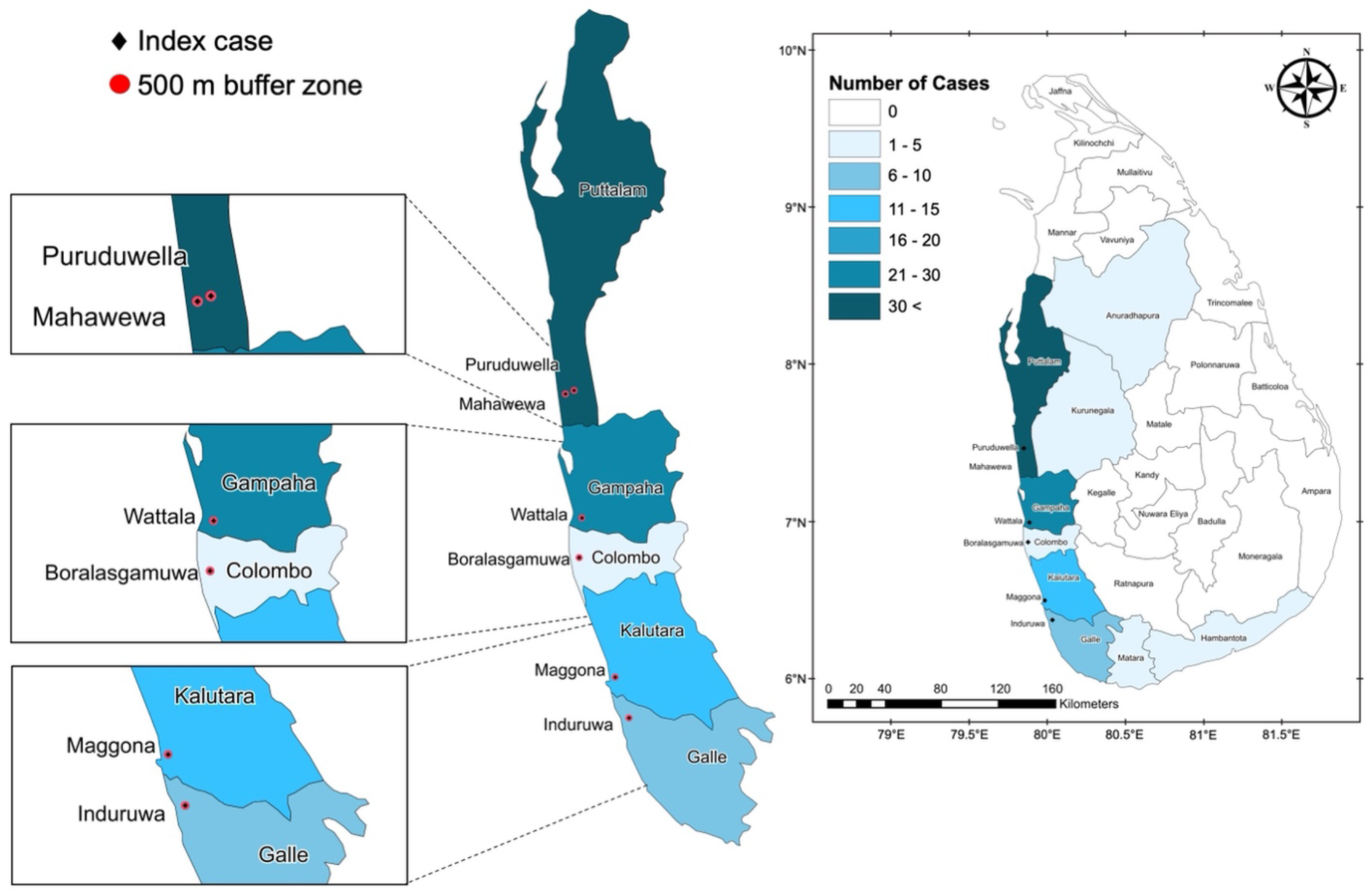
Distribution of brugian filariasis cases in Sri Lanka and sample collection sites. Districts are colored by decreasing intensity, according to the number of human brugian filariasis cases reported from each district from 2006 to 2021. The collection sites were the most recent cases of human brugian filariasis in each district at the time of the study, marked by a black diamond, with the buffer zone marked in red; Puruduwella-Puttalam District (S_1_), Maggona-Kalutara District (S_2_), Wattala-Gampaha District (S_3_), Induruwa-Galle District (S_4_), Boralesgamuwa-Colombo District (S_5_), and Mahawewa-Puttalam District (S_6_).

Blood samples were collected during the daytime from ear-lobe pricks of domestic and stray animals (dogs and cats) within the study area. TBSs were prepared, stained with Giemsa, and microscopically examined for *Brugia* sp. infections. DNA was extracted as described above. Mosquitoes were collected using dog-baited, gravid, and window traps at each study site as described previously [24]. The head and thorax regions of the collected mosquitoes were dissected and observed under a stereomicroscope for the presence of the infective L3 stage of the *Brugia* sp. DNA was extracted from the head and thorax of potentially infective *Brugia* sp. DNA was extracted from mosquitoes with visible larvae using the methods described above.

### Sequence analysis of *Brugia* sp. from infected animals and potentially infective mosquitoes

The *SLXI*, *COXI,* and HhaI regions were amplified from parasites extracted from potentially infective mosquitoes and infected animals, as mentioned above, and then sequenced. DNA sequences of the *SLXI*, *COXI,* and HhaI regions of the brugian parasites found within humans, animals, and mosquitoes, and of the FR3 strains of *B. malayi* and *B. pahangi* were aligned using CLC Genomic Workbench v23.0.1.

### Blood meal analysis of *Brugia* sp. infected mosquitoes

To determine recent feeding by potentially infected mosquitoes, the human, cat, and dog DNA was amplified from the thoraces. The 272 bp and 659 bp mitochondrial Cytochrome b region of humans and dogs were amplified using the universal reverse primer UNVR; 5’-AGTGGGYGRAATATTATGC-3’ and HMNF; 5’-CTCGGCTTACTT CTCTTCC-3’, and DOGF; 5’-AGCCTATATTACGGATCCTATG-3’ respectively [29]. The PCR mixture was heated for 5 min at 95 °C, followed by 12 cycles of 94 °C for 30 s, 57 °C for 30 s and 72 °C for 50 s. Two additional sets of 12 cycles each were performed, using decreasing annealing temperatures of 56 and 55 °C respectively. A final elongation at 72 °C was performed for 5 min.

## Results

### Physical characteristics of the prevalent Sri Lankan brugian parasite are consistent with *B. malayi*

Morphological identification of parasites present in human blood samples was based on Giemsa-staining patterns of TBSs and worm body measurements. Giemsa-stained TBSs from fourteen (n=14) human blood samples were observed for the presence of *Brugia* sp. parasites. A pink sheath, a visible cephalic space, loosely arranged body nuclei, and two terminal and sub-terminal nuclei characteristic of *Brugia* microfilariae were observed in TBSs [Fig 2 (A)]. The body measurements of the *Brugia* sp. recorded in this study differed from those of previously reported isolates in Sri Lanka; however, the measurements are within the range reported for *B. malayi* from other regions in South and Southeast Asia, as evident by the Principal Component Analysis [Table 1, Fig 2(B)].

**Fig 2.**
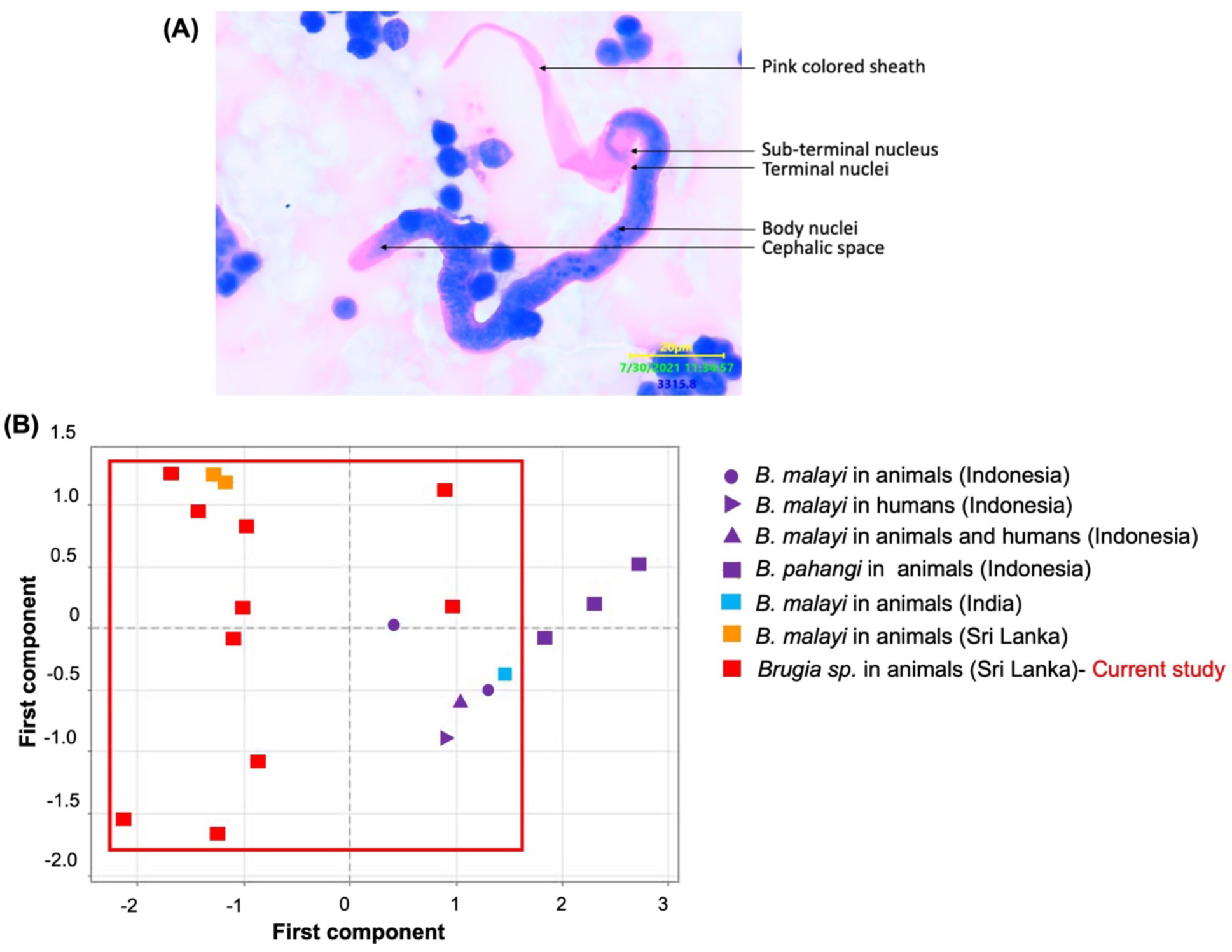
Morphological and morphometric analysis revealed similarity to *B. malayi*. (A) Microfilariae of *Brugia* sp. found in Giemsa-stained TBSs of infected humans. (B) Morphometric analysis of the *Brugia* sp. is within the range of previously identified *B. malayi*. The region enclosed in the red square indicates the range of body measurements for *B. malayi*.

**Table 1.**
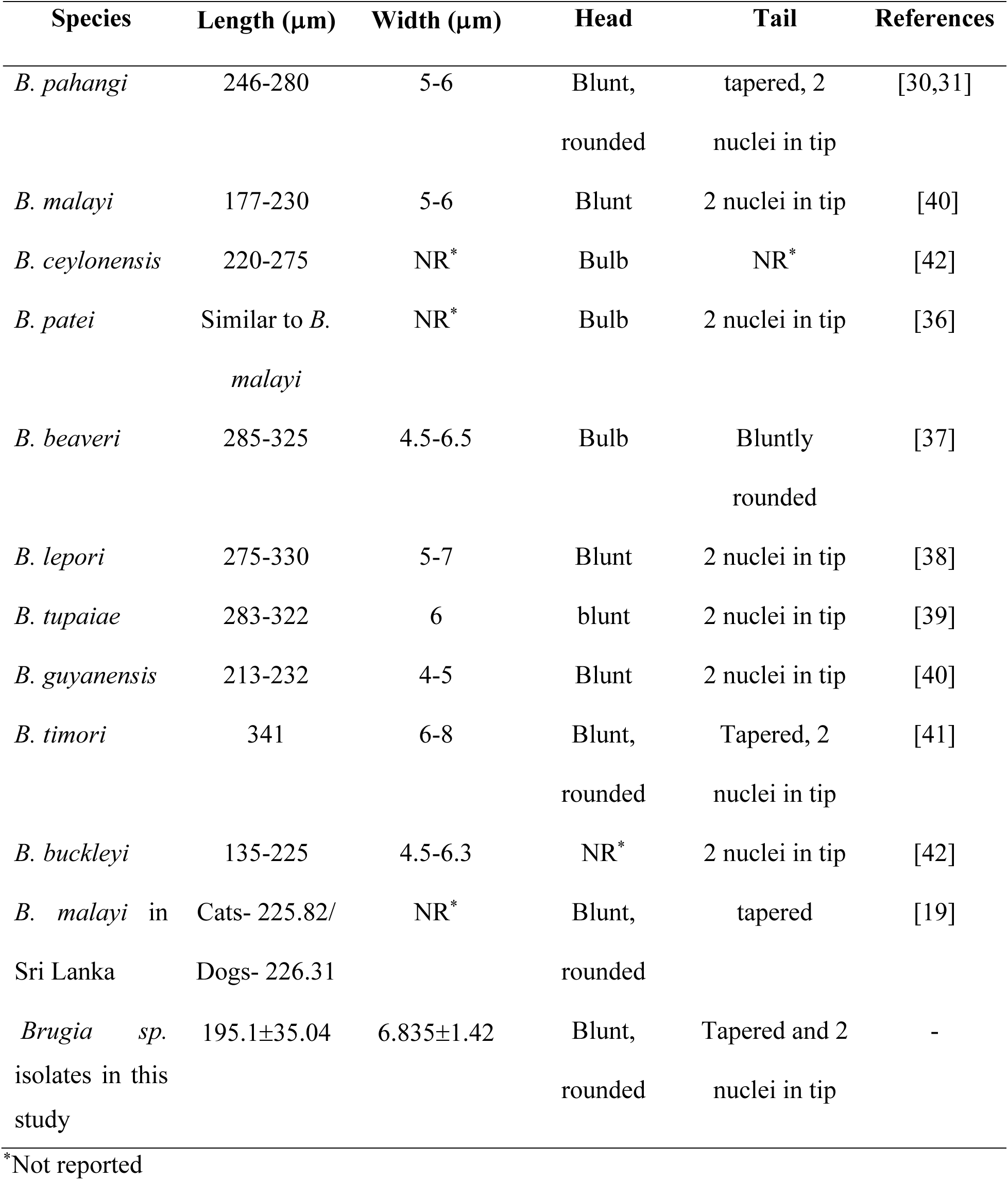
Morphometric comparison of the *Brugia* sp. in Sri Lanka with other published data.

| Species | Length (μm) | Width (μm) | Head | Tail | References |
| --- | --- | --- | --- | --- | --- |
| *B. pahangi* | 246-280 | 5-6 | Blunt,<br>rounded | tapered, 2<br>nuclei in tip | [30,31] |
| *B. malayi* | 177-230 | 5-6 | Blunt | 2 nuclei in tip | [40] |
| *B. ceylonensis* | 220-275 | NR* | Bulb | NR* | [42] |
| *B. patei* | Similar to *B. malayi* | NR* | Bulb | 2 nuclei in tip | [36] |
| *B. beaveri* | 285-325 | 4.5-6.5 | Bulb | Bluntly<br>rounded | [37] |
| *B. lepori* | 275-330 | 5-7 | Blunt | 2 nuclei in tip | [38] |
| *B. tupaiae* | 283-322 | 6 | blunt | 2 nuclei in tip | [39] |
| *B. guyanensis* | 213-232 | 4-5 | Blunt | 2 nuclei in tip | [40] |
| *B. timori* | 341 | 6-8 | Blunt,<br>rounded | Tapered, 2<br>nuclei in tip | [41] |
| *B. buckleyi* | 135-225 | 4.5-6.3 | NR* | 2 nuclei in tip | [42] |
| *B. malayi* in<br>Sri Lanka | Cats- 225.82/<br>Dogs- 226.31 | NR* | Blunt,<br>rounded | tapered | [19] |
| *Brugia* sp.<br>isolates in this<br>study | 195.1±35.04 | 6.835±1.42 | Blunt,<br>rounded | Tapered and 2<br>nuclei in tip | - |

\*Not reported

### Molecular and phylogenetic analysis of *Brugia* sp. infecting humans in Sri Lanka reveals distinction from *B. malayi* and *B. pahangi*

#### Molecular analysis

Fourteen *Brugia sp.* positive human blood samples were subjected to homology analysis using partial *COXΙ* (460 bp/517 bp), partial HhaΙ (239 bp/322 bp), and partial *SLXΙ* (230 bp/294 bp) to assess molecular similarities among the *Brugia* sp. in different districts in Sri Lanka. The multiple sequence alignment of the *COXI* region was found to be identical across all infected human blood samples, while MSA of the HhaΙ and *SLXI* regions identified two variant sequences each (Fig 3).

**Fig 3.**
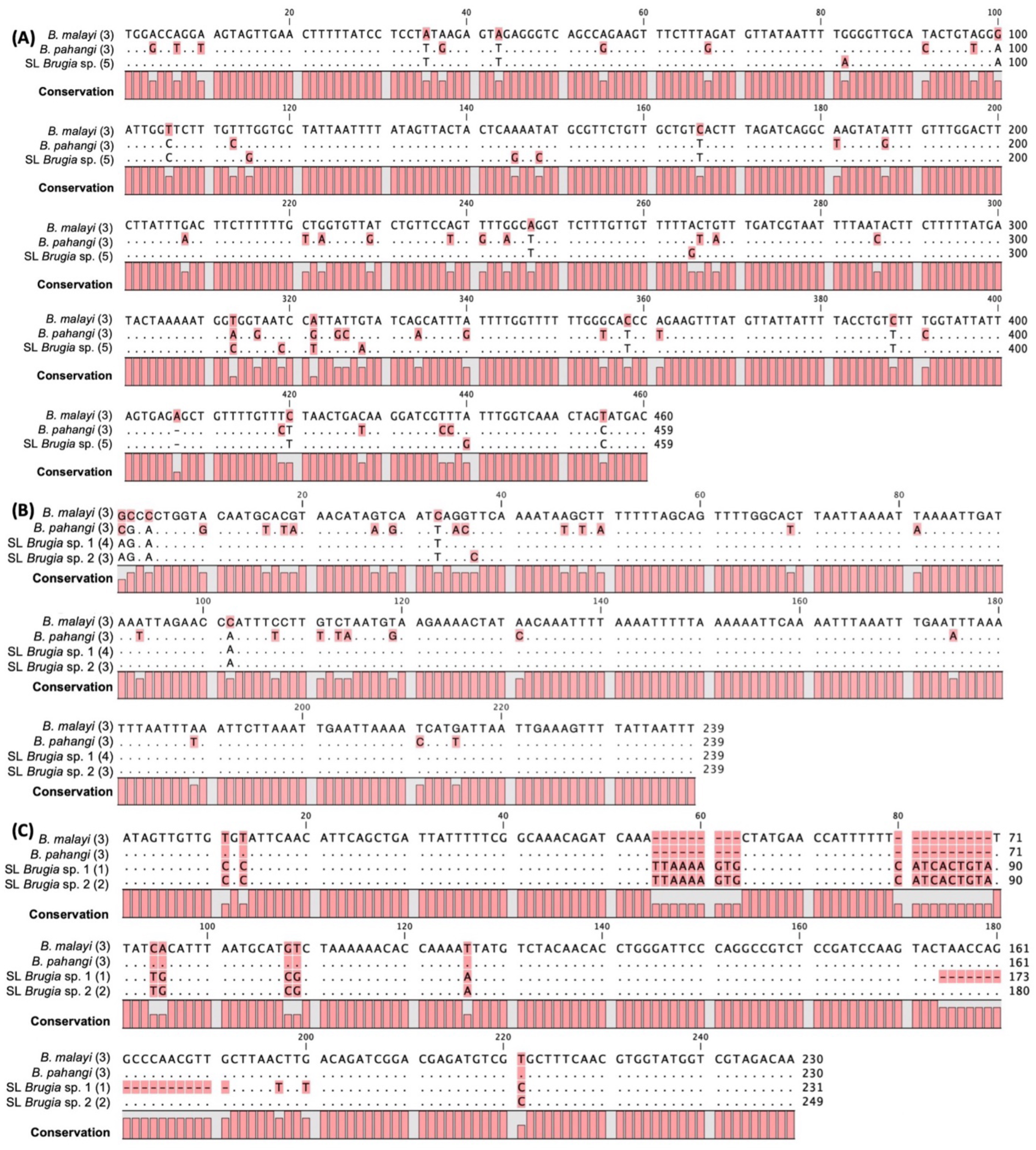
Multiple sequence alignment of three genomic regions of the *Brugia* sp. isolated in this study showed a closer similarity to *B. malayi* than *B. pahangi.* Multiple sequence alignment of (A) *COXΙ* region, (B) HhaΙ region and (C) *SLXΙ* region. The consensus sequence for the FR3 *B. malayi* is shown as the reference, with conserved residues in *B. pahangi* and the *Brugia* sp. in Sri Lanka represented by dots and changes highlighted in pink. The number of samples used for each of the sequences is detailed in parentheses.

The *COXΙ* region analysis identified the same sequence in all field-collected infected human samples (n=5). Homology analysis of the partial *COXI* region (460 bp) revealed 95.4% and 90.0% similarity to FR3 strains of *B. malayi* and *B. pahangi*, respectively, with twenty (20) SNPs and one (01) deletion in comparison to *B. malayi* and forty-three (43) SNPs and one deletion in comparison to *B. pahangi* [Fig 3(A)]. The HhaI region (239 bp) alignment identified 2 variant sequences differed by one SNP between cytosine (C) (n=3) and thymine (T) (n=4) at 37 bp. However, the field collected *B. malayi* showed 97.5-97.9% similarity to the FR3 strains of *B. malayi* [five (05) SNPs], and 88.1% similarity to the FR3 strains of *B. pahangi* [twenty-six (26) SNPs and a one (01) bp insertion] [Fig 3(B)]. Sequence alignment of the partial *SLXI* region revealed two variant parasite sequences in humans (n=3), characterized by an 18 bp difference at positions 174-191 bp and two single-nucleotide differences at positions 197 and 200. In comparison to the brugian FR3 strains, a 9 bp insertion was observed at 55-63 bp, a 10 bp addition was observed at 80-89, while 8 SNPs were observed in both sequence types at positions 11, 13, 94, 95, 108, 109, 126, and 221 [Fig 3(C)].

#### Phylogenetic analysis

Bayesian phylogenetic analysis of the mitochondrial *COXΙ* region revealed that *Brugia* sp. forms a significant, polyphyletic, well-supported clade with a posterior probability of 1 [Fig 4(A)]. Representative *COXΙ* sequences from infected human samples were from Puttalam (2019, n=1 and 2021, n=1), Galle (2022, n=1), Gampaha (2022, n=1), and Kalutara (2022, n=1). The *Brugia* sp. in Sri Lanka forms a polyphyletic group with *B. pahangi* sequences, which clearly separates from *B. malayi* sequences reported from Thailand with a posterior probability of 0.6366. This *Brugia* sp. also clearly separates from *B. pahangi* sequences reported from Thailand and Malaysia with a posterior probability of 0.8644. Notably, the Sri Lankan isolates cluster near *Brugia* sp. sequences reported from dogs in India, and with previously reported *Brugia* sp. from Sri Lanka.

**Fig 4.**
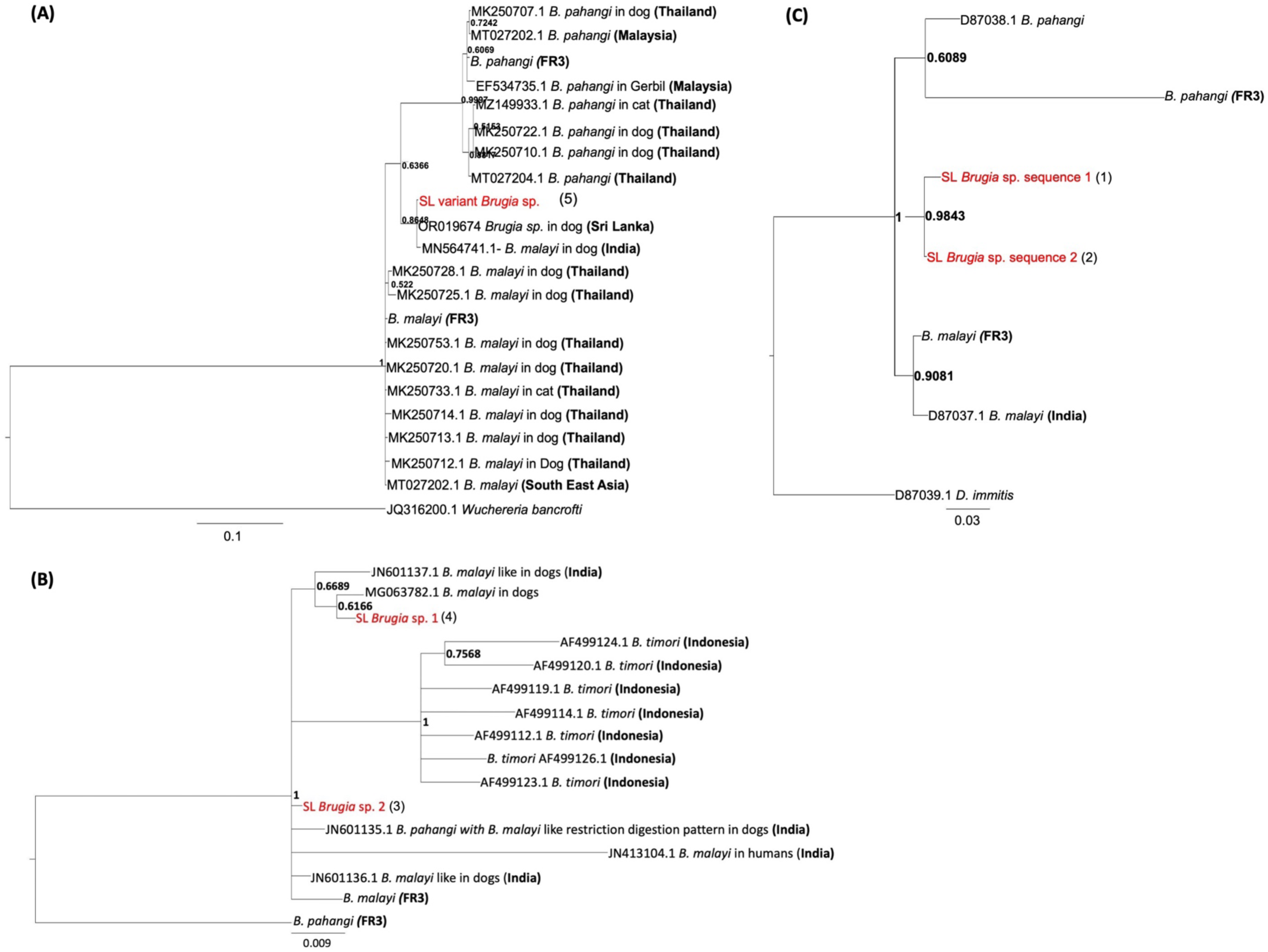
Phylogenetic analysis of (A) *COXI* region, (B) HhaI region, and (C) *SLXI* region reveals the prevalent brugian parasite in Sri Lanka is similar to, but distinct from, *B. malayi* parasites isolated from South and Southeast Asia. The posterior probability values are shown at the branch nodes. Sequences retrieved from GenBank and published studies are labeled with accession numbers and corresponding references. All FR3 sequences were sequenced in this study and confirmed with at least 3 samples. The number of human samples used for the *Brugia* sp. in Sri Lanka is detailed in parentheses.

In contrast, Bayesian phylogenetic analysis of the HhaI region from Puttalam (2020, n=3; 2019, n=1), Kalutara (2019, n=2), and Galle (2020, n=1) revealed that the *Brugia* sp. detected in infected humans in Sri Lanka clusters within the *B. malayi* lineage [Fig 4(B)]. The Sri Lankan *Brugia* sp. group is closely related to the reference *B. malayi* FR3 strain and to *B. malayi* and *B. malayi*-like sequences reported from dogs and humans in India, forming a well-supported monophyletic group. These isolates are clearly separated from *B. pahangi* FR3 reference sequences, with a posterior probability of 1, indicating a closer phylogenetic affinity to *B. malayi* than to *B. pahangi*. Furthermore, the Sri Lankan isolates are distinct from *B. timori* sequences from Indonesia, with a posterior probability of 1, forming a polyphyletic group that supports the placement of the Sri Lankan *Brugia* sp. within the *B. malayi* complex.

Phylogenetic analysis of the two *SLXΙ* sequences showed that the *Brugia* sp. in Sri Lanka forms a polyphyletic group with FR3 strains of *B. malayi* and *B. pahangi*, and is clearly separated from both FR3 strains [Fig 4(C)]. The two identified sequences are significantly different from each other, with a posterior probability of 0.9843. Representative *SLXΙ* sequence 1 was from a human blood sample from Puttalam (2021, n=1), and *SLXΙ* sequence 2 human samples were from Gampaha (2020, n=1) and Kalutara (2019, n=1).

The mean K2P genetic divergence analysis of the mitochondrial *COXΙ* region showed less than 3% divergence between the *Brugia* sp. identified in the present study and previously reported *Brugia* sp. from Sri Lanka, as well as *B. malayi* isolates from India (Table 2). In contrast, divergence values exceeding 3% were observed when compared with *B. malayi* isolates from Thailand. These findings suggest that the brugian parasite characterized in this study is genetically closer to the South Asian *B. malayi* reported from Sri Lanka and India, while showing greater differentiation from Thai *B. malayi* isolates.

**Table 2.** Kimura 2-parameter (K2P) genetic divergence analysis using the *COXΙ* region between the *Brugia* sp. in Sri Lanka and other brugian parasites indicates a closer relationship to *B. malayi* in dogs from India and distinct from other parasites.

| Species | NCBI accession number | Interspecific genetic distances with the Sri Lankan *Brugia* sp. identified in this study |
| --- | --- | --- |
| *B. pahangi* in dogs (Thailand) | MK250707.1 | 9.84 |
| *B. pahangi* (Malaysia) | MT027202.1 | 9.84 |
| *B. pahangi* (FR3)* |  | 10.60 |
| *B. pahangi* in gerbil (Malaysia) | EF534735.1 | 10.87 |
| *B. pahangi* in cats (Thailand) | MZ149933.1 | 10.37 |
| *B. pahangi* in dogs (Thailand) | MK250722.1 | 10.35 |
| *B. pahangi* in dogs (Thailand) | MK250710.1 | 10.62 |
| *B. pahangi* (Thailand) | MT027204.1 | 10.10 |
| *B. malayi* in dogs (India) | MN564741.1 | 0.45 |
| *B. malayi* in dogs (Thailand) | MK250728.1 | 4.63 |
| *B. malayi* in dogs (Thailand) | MK250725.1 | 5.36 |
| *B. malayi* (FR3)* |  | 4.38 |
| *B. malayi* in dogs (Thailand) | MK250753.1 | 4.38 |
| *B. malayi* in dogs (Thailand) | MK250720.1 | 4.14 |
| *B. malayi* in dogs (Thailand) | MK250733.1 | 4.14 |
| *B. malayi* in dogs (Thailand) | MK250714.1 | 4.87 |
| *B. malayi* in dogs (Thailand) | MK250713.1 | 4.14 |
| *B. malayi* in dogs (Thailand) | MK250712.1 | 4.63 |
| *B. malayi* in dogs (South East Asia) | MT027202.1 | 4.14 |
| *W. bancrofti* | JQ316200.1 | 75.14 |

\*Sequenced in this study

Furthermore, as shown in Table 2, higher K2P divergence values were observed in comparison with *B. pahangi*. Overall, pairwise K2P distances ranged from 4–6% when compared with *B. malayi* sequences and from 9–11% when compared with *B. pahangi* sequences, indicating marked genetic separation from *B. pahangi* and moderate divergence within *B. malayi* lineages.

### Integrated analysis of infected animals, humans, and potentially infective mosquitoes provides evidence for the zoonotic potential of the Sri Lankan *Brugia* sp

A total of one hundred and twenty-four (124: n_dog_=84, n_cat_=28, n_cattle_=12) domestic and stray animals from six study sites across southwest Sri Lanka [S_1_ (n=39), S_2_ (n=28), S_3_ (n=12), S_4_ (n=15), S_5_ (n=12), and S_6_ (n=18)] were screened for *Brugia* sp. infections using Giemsa-stained TBS observations [Fig 5(A)]. *Brugia* sp. and *D. repens* were identified based on characteristic morphological features. *Brugia* sp. was identified as described earlier, and *D. repens* by its hook-shaped caudal region, rounded cephalic region, long cephalic space with two nuclei, and body nuclei that do not extend to the tip of the tail. Both *Brugia* sp. and *D. repens* were detected in TBS from the S_1_ site (*Brugia* sp., 14/39; 36%; *D. repens,* 24/39, 62%), only *D. repens* from the S_2_ site (11/28, 39%), only *Brugia* sp. from the S_3_ site (2/12, 16.6%), and no brugian parasites from the S_4_, S_5,_ and S_6_ sites [Fig 5(A)]. All TBS positive for *Brugia* sp. infections were confirmed by *Brugia* species-specific HhaΙ PCR analysis.

**Fig 5.**
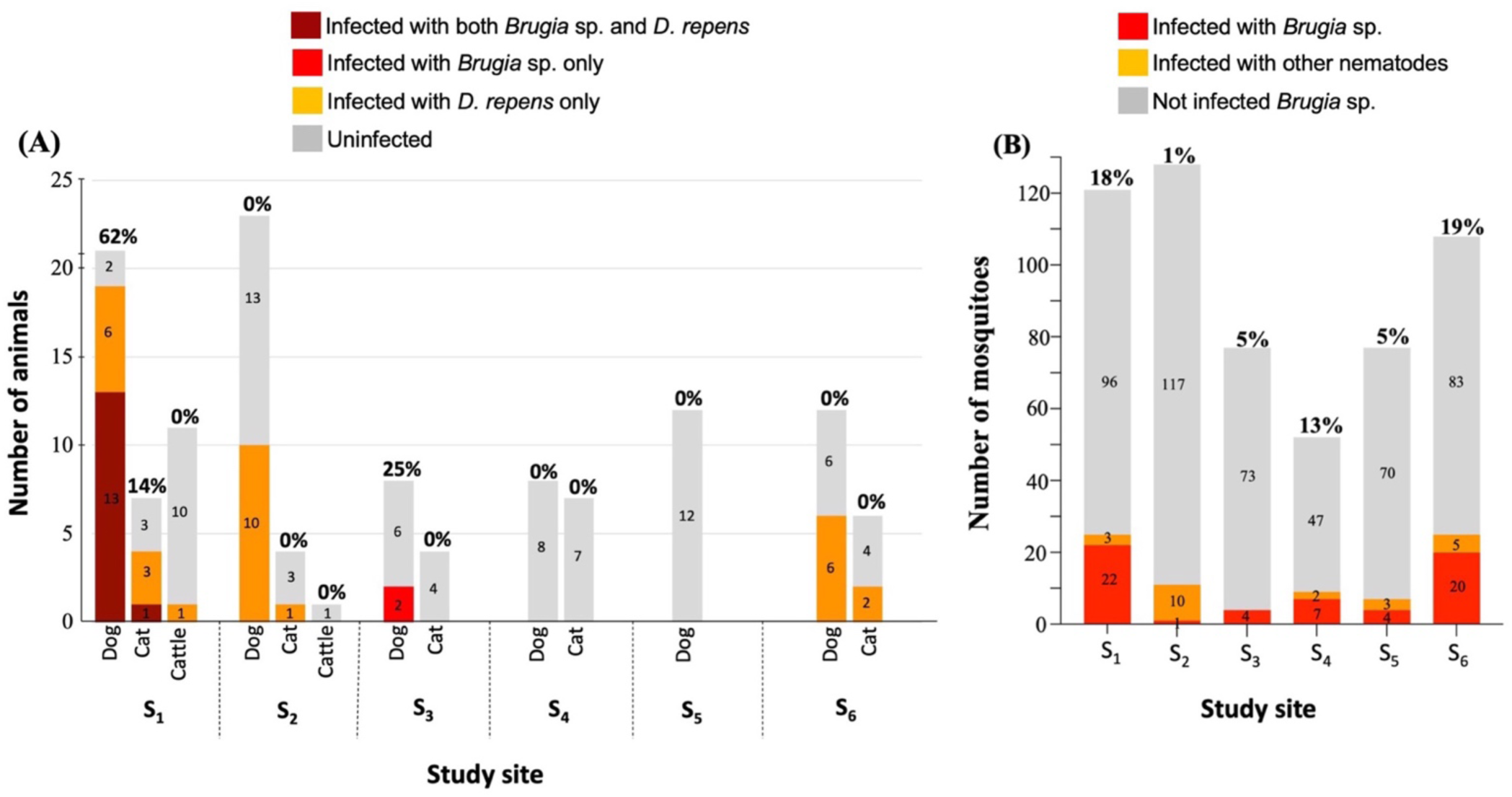
Brugian infections were identified within (A) animals and (B) mosquitoes near human index cases. **(A)** Number of animals infected with *Brugia* sp. as determined by thick blood smears (TBSs). Percentage values of brugian infections are shown above each bar and the proportion of TBS-positive animals from all animals surveyed at each site are indicated in marron, orange and red. **(B)** Number of mosquitoes with brugian parasites identified by dissection and confirmed using *Brugia* species-specific PCR. Percentage values are displayed above each bar and the proportion of dissected mosquitoes subsequently confirmed by HhaΙ PCR are indicated in red and orange. Puruduwella-Puttalam District (S_1_), Maggona-Kalutara District (S_2_), Wattala-Gampaha District (S_3_), Induruwa-Galle District (S_4_), Boralesgamuwa-Colombo District (S_5_) and Mahawewa-Puttalam District (S_6_).

A total of seven hundred sixty-six (766) mosquitoes from six study sites were analyzed for *Brugia* sp. infections by dissecting the head and thorax regions together and confirmed by *Brugia* species-specific HhaΙ PCR [Fig 5 (B)]. Nematodes were observed in the head and thorax regions of 125 mosquito species: S_1_ (47/133, 57.9%), S_2_ (29/214, 13.6%), S_3_ (4/151, 2.6%), S_4_ (9/59, 15.2%), S_5_ (11/81, 13.6%), and S_6_ (25/128, 19.5%). Of all the dissection-positive mosquitoes 61.6% (n=77) were identified as potentially infective *Brugia* sp. due to the presence of the L3 larval stage of *Brugia* sp.; 27.1% (36/133) from S_1_, 5.6% (12/214) from S_2_, 1.3% (2/151) from S_3_, 5.1% (3/59) from S_4_, 4.9% (4/81) from S_5,_ and 3.6% (20/128) from S_6._

Body measurements of the brugian parasite identified in infected cats and dogs differed from those of the variant *Brugia* sp. previously reported in Sri Lanka, but were similar to those of the prevailing *Brugia* sp. in infected humans, with measurements within the range for *B. malayi*. The mean total body length was smaller than that reported for *B. malayi* in Sri Lanka (Table 3). The cephalic space exhibited increased dimensions in both length and width; however, the width-to-length ratio was lower than in earlier reports.

**Table 3.** Comparison of the body measurements of microfilaria of *B. malayi* in infected animals’ blood in the current study and previously reported *B. malayi* in Sri Lanka.

| Parasite | Animal | Mean total body length (µm) | Mean length of cephalic space (µm) | Mean width of cephalic space (µm) | Cephalic space width: length ratio | References |
| --- | --- | --- | --- | --- | --- | --- |
| *B. malayi* | Dog | 225.82 | 5.56 | 4.87 | 0.86 | [19] |
|  | Cat | 226.31 | 5.47 | 4.84 | 0.88 |  |
| *Brugia* sp. isolates in this study | All together | 195.1±35.04 | 6.687±1.68 | 5.22±0.52 |  | - |
|  | Dog | 195.3±36.82 | 6.414±1.69 | 5.176±0.51 | 0.67 |  |
|  | Cat | 194.1±36.03 | 8.050±0.92 | 5.435±0.69 |  |  |

Homology analysis of the *COXI* region of *Brugia* sp. from infected humans, animals, and mosquitoes revealed a single conserved sequence across humans, animals, and mosquitoes [Fig 6 (A)]. In contrast, analysis of the HhaΙ region identified 2 sequences, one of which is shared among humans, animals, and mosquitoes, and the other shared only among humans and mosquitoes [Fig 6(B)]. Further, analysis of the *SLXΙ* region identified three distinct sequences: one shared among humans, animals, and mosquitoes, one detected exclusively in a human sample, and another detected only in a mosquito sample [Fig 6(C)]. The presence of the same brugian parasite in humans, animals, and mosquitoes suggests the possibility of transmission between humans and animals via mosquitoes.

**Fig 6.**
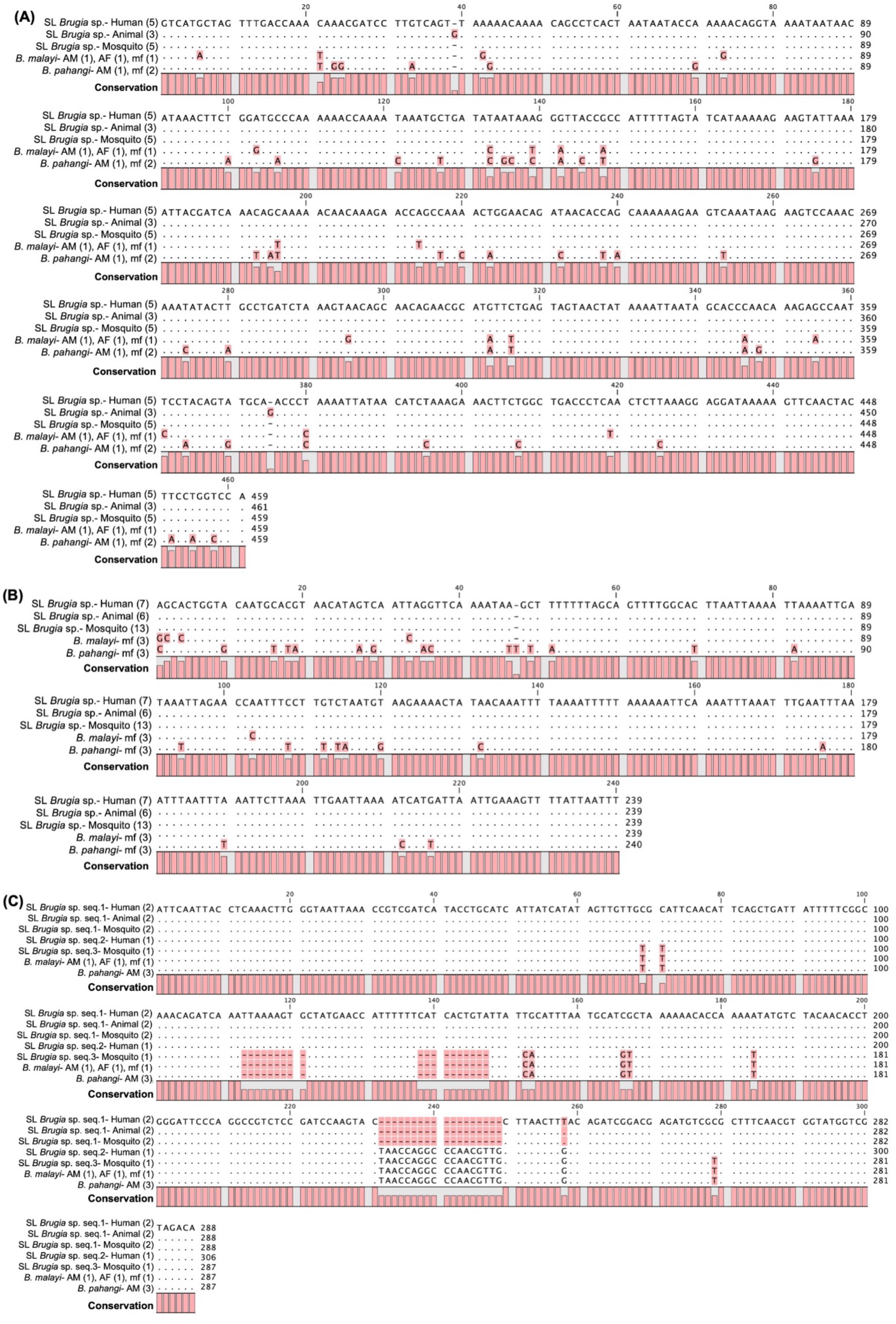
Multiple sequence alignment of different genomic regions of *Brugia* sp. isolated from humans, animals, and mosquitoes in Sri Lanka identified shared sequences among hosts and vectors, suggesting the potential of zoonosis. The (A) *COXΙ* region, (B) HhaΙ region, and (C) *SLXΙ* region of samples isolated from humans, animals, and mosquitoes were aligned with sequences obtained separately from different life stages of *B. malayi* and *B. pahangi*. Life stages from which sequences were obtained are denoted within parentheses: AM-adult male, AF-adult female, mf-microfilariae. Conserved residues are shown by dots, and variable bases are highlighted in pink. The number of samples used to generate consensus sequences is detailed in parentheses.

Mosquitoes harboring *Brugia* sp. L3 larval stage in the head and thorax regions were considered as potentially infective because they can support the development of *Brugia* sp. parasites from microfilariae to the infective L3 stage. To investigate the possibility of transmission of these parasites between humans and animals, the blood meals of potentially infective mosquitoes were analyzed for human and animal (dog and cat) blood. Of the 794 dissected mosquitoes, 77 were identified as potentially infective; *Ma. annulifera* (n=3/77, 3.9%), *Ma. indiana* (n=19/77, 24.7%), *Ma. uniformis* (n=11/77, 14.3%), *Cx. lophoceraomyia* (n=2/77, 2.6%), *Cx. tritaeniorhynchus* (n=22/77, 28.6%), *Cx. quinquefasciatus* (n=6/77, 7.8%), *Cx. vishnui* (n=2/77, 2.6%), *Ar. subalbatus* (n=10/77, 13%) and *Cq. crassipes* (n=2/77, 2.6%). Of those, both human and animal blood were detected in 28.6% of the mosquitoes (n=22/77); *Ma. annulifera* (n=1/3), *Ma. indiana* (n=7/19), *Cx. tritaeniorhynchus* (n=10/22), *Cx. quinquefasciatus* (n=2/6), *Ar. subalbatus* (n=1/10) and *Cq. crassipes* (n=1/2), suggestive of the potential for zoonotic transmission of the prevailing *Brugia* sp. with *Ma. indiana* and *Cx. tritaeniorhynchus* having the highest vector potentiality of zoonotic transmission (Fig 7).

**Fig 7.**
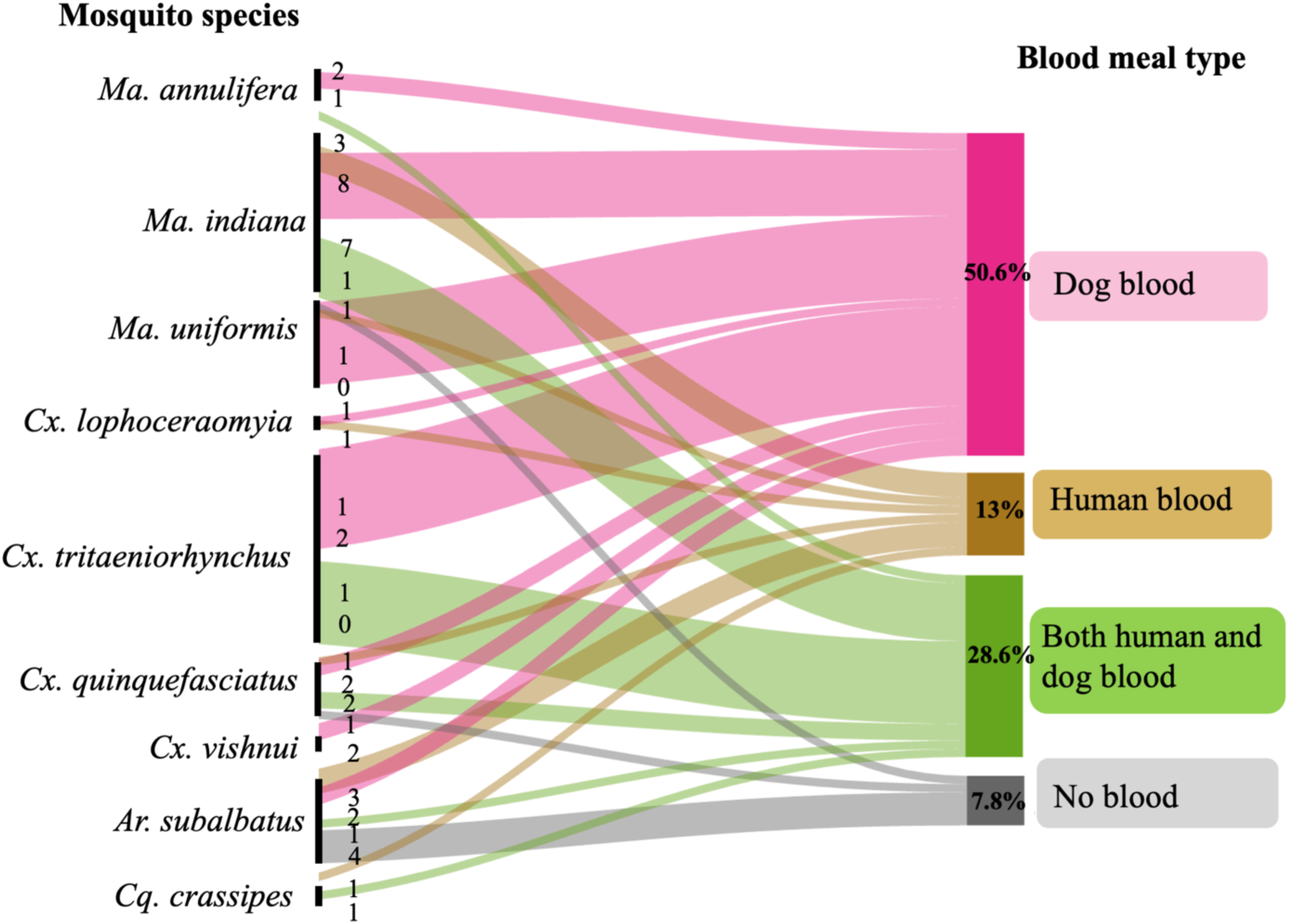
Potentially infective mosquitoes show blood meals from both animals and humans, suggesting potential zoonosis of the prevailing *Brugia* sp. The thickness of links in the Sankey diagram is proportional to the total abundance of mosquitoes with that specific blood meal type across the sample.

Further, previous research has identified three (3) SNPs in the mitochondrial genome at nucleotide positions 2511, 2514, and 3130 to deduce the origin of *B. malayi* parasites [43]. Sequences from a total of 15 samples of the brugian parasite collected from animals, humans, and mosquitoes were compared with those of the *B. malayi* FR3 strain and with GenBank sequences from Thailand (Fig 8).

**Fig 8.**
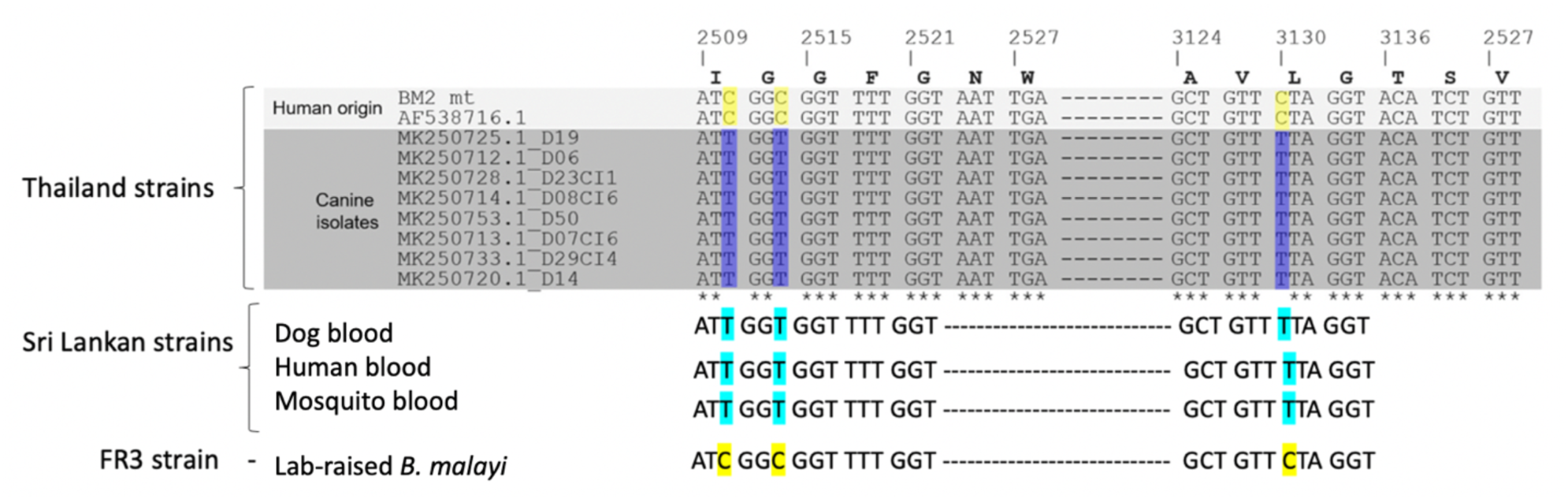
Single Nucleotide Polymorphism analysis revealed a zoonotic origin of the *Brugia* sp. in Sri Lanka. The *COXΙ* region between 2509-2527 obtained from *Brugia* sp. dogs, humans, mosquitoes, and *B. malayi* obtained from FR3 were aligned with those from *B. malayi* Thailand strains retrieved from GenBank using CLC Genomic Main Workbench. The C/T SNPs at positions 2511, 2514, and 3130 could distinguish *B. malayi* isolates as either of human or canine origin.

## Discussion

Lymphatic filariasis (LF) is considered a “potentially eradicable” disease by the World Health Organization [44]. However, post-validation surveillance and recent research suggest the reemergence of brugian filariasis within Sri Lanka [16,17,19]. Recent studies show that the prevailing brugian parasite differs from the previously reported one, with an expanded range of potential vector mosquito species [24], and may possess a zoonotic transmission potential. However, knowledge regarding this brugian parasite is limited, and this study aimed at characterizing the prevailing brugian parasite in Sri Lanka to better inform effective control and prevention measures. Here we report that the prevailing brugian parasite in Sri Lanka bears physical characteristics of *B. malayi* in agreement with previous reports [19], with body measurements within the range of those of *B. malayi,* yet different from those previously reported from Sri Lanka, India, Indonesia, and Thailand [19,45–47]. Although morphological observations facilitate identification of the brugian parasite to the genus level, differentiation at the species level requires molecular analysis [48].

This study provides sequence information of *SLXI* and HhaI regions of brugian parasites found in humans, animals, and mosquitoes, and *COXI* in mosquitoes in Sri Lanka for the first time. All three genomic regions used for molecular and phylogenetic analysis in this study are well recognized for investigating genetic variability. *SLXΙ* is a widely analyzed region of *Brugia* sp. for the molecular identification of the parasite [20,49]. The mitochondrial genome (*COXΙ*) is used to identify genetic variability and phylogenetic relationships, since it evolves more rapidly than the nuclear genome [50,51]. The use of HhaΙ sequences in distinguishing taxonomic relationships at the DNA level is promising, since it doesn’t code for any protein or RNA product and therefore, is able to evolve much faster than the coding sequences [52]. Further, the HhaΙ repetitive sequence is *Brugia* species-specific and thus cannot be found in other closely related species, such as *W. bancrofti* [53].

Characterization of the brugian parasite in humans in Sri Lanka via sequence homology and phylogenetic analyses based on *COXI,* HhaI, and *SLXI* regions revealed that the brugian parasite in Sri Lanka is distinct from FR3 reference strains of both *B. malayi* and *B. pahangi.* This could be due to mutations accumulated over time in the FR3 strains since they have been maintained in Mongolian Gerbils (*Meriones unguiculatus*) for more than 15 years [54]. To circumvent this, sequence information of brugian parasites in Sri Lanka and the Southeast Asian region were used from the NCBI database for a more representative understanding of the phylogenetic relationship of these parasites.

*COXΙ* phylogeny showed clustering with a brugian parasite previously reported from a dog in Sri Lanka and India, in agreement with recent literature [22,23]. Phylogenetic analysis of the HhaⅠ region revealed a close affinity to isolates reported from India, commonly referred to as *B. malayi*-like parasites [55]. Notably, this is the first documentation of HhaI sequences of these parasites in Sri Lanka, and the lack of previous records prevents historical comparison. Further, the lack of availability of HhaI sequences other than those of *B. malayi* from India and *B. timori* from Indonesia prevents a comprehensive analysis as with *COXI*. Further, this is the first report of *SLXI* sequences from Sri Lanka. This highlights the need for more comprehensive molecular characterization of these parasites regionally to accurately delineate the taxonomic status and epidemiological origin.

Genetic divergence analysis supports the phylogenetic analyses’ findings of *COXI* and HhaI regions, indicating that the *Brugia* sp. identified in this study is genetically closer to South Asian *B. malayi* isolates reported from Sri Lanka and India, while showing greater divergence from Thai *B. malayi* isolates. However, the lack of genetic information on *B. ceylonensis,* the previously identified brugian parasite in Sri Lanka [56], prevents historical comparisons of the prevailing parasite and remains a limitation for the current study.

Confirmation of the zoonotic potential of parasites is imperative to achieve effective brugian filariasis control within the country. Here we provide identical sequences of the brugian parasite across humans, animals, and mosquitoes for the first time in Sri Lanka. This, together with bloodmeals comprising both human and animal blood within infective mosquitoes, provides concrete molecular evidence of the zoonotic transmission potential of this parasite. Previous reports in India have reported *B. malayi* and *B. malayi-*like parasite infections in dogs and cats, suggesting the possibility of zoonotic transmission [17,20,49]. Mallawarachchi *et al* predicted the zoonotic nature of the brugian parasite in Sri Lanka based on its subperiodic behavior [16], and Gunaratna *et al.* further contributed support of zoonosis by reporting monophyletic clustering of *COXI* sequences of animals and humans [22], whether these parasites had evolved separately within the two hosts remained unanswered. However, the present study provides the first comprehensive evidence supporting the zoonotic potential of the brugian parasite in Sri Lanka by integrating data from infected humans, animal hosts, and vector mosquitoes, using three independent genomic regions and bloodmeal analysis of infective mosquitoes. These findings underscore the importance of the One Health approach to understanding transmission dynamics and to designing effective control strategies for brugian filariasis.

Of further interest are the sequence variants of the three regions identified in this study. Of the two variant sequences of HhaI, one was present in humans, animals, and mosquitoes, while the other was present only in humans and mosquitoes. Of the three sequence variants of *SLXI* one type was found in all hosts and vectors, while one was found only in humans, and the other only in mosquitoes. Interestingly, Gunaratna *et al.* reported two human and one animal *COXI* sequences, one of which is shared between humans and animals yet differs by one base pair from ours [22]. These changes could represent a parasite that has evolved over time to cross the species barrier, developing adaptive mutations for persistent existence in the new host. This conjuncture of the parasite could support the recent identification of a wide range of potential vector mosquito species of it [24]. This possibility is alarming and warrants the need for region-specific genetic data on the variability of this parasite across geographic locations to better understand the current situation, to prevent the spread and establishment of a novel genetic variant of the brugian parasite.

Interestingly, comparison of the C*OXΙ* sequence data with a study conducted in Thailand identified key SNPs (Single Nucleotide Polymorphisms) that suggest a canine origin of the brugian parasite in Sri Lanka [43]. Identification of the canine-originated brugia strain in humans, animals, and mosquitoes further suggests the risk of potential zoo-anthroponoses of this parasite in Sri Lanka. According to the findings of the current study, the reemergence of the disease after a few decades of quiescence can be explained as follows. When Sri Lanka received the certificate of elimination of LF in 2016, the microfilaria prevalence rate was 0.05%, in accordance with the criterion stipulated by WHO for verification of elimination [15]. However, it is possible that at the time the certificate was received, the parasitic agent was still present in the canine and feline populations, which were not analyzed. Furthermore, due to genetic variation within *B. malayi*, the parasite may have found its way back into the human population, as evidenced by the continued reporting of an increasing number of brugian filariasis patients even after the certificate of elimination was issued [57,58]. Alarmingly, the number of reported brugian filariasis cases currently exceeds that of bancroftian filariasis, a reversal of the previously observed trend of higher bancroftian cases than those of brugian cases.

Careful screening and surveillance of at-risk populations, including individuals with recent travel or migration history from endemic regions, may be warranted to prevent the potential introduction or spread of the parasite. Here, not all animals in the area could be surveyed; therefore, only an estimate of the disease burden can be made. The derived prevalence rates are likely to underestimate the true prevalence due to the relatively low sensitivity of the screening tool (Giemsa-stained TBS observation). Also, although the results suggest zoonosis, the direction of zoonosis, whether it is anthropozoonosis, zooanthropozoonosis, or transferred in both directions, or whether it was initially transmitted from animals to humans and has now established itself in humans and is no longer transmitted back to animals, is not established.

Results from the current study add a layer of complexity to the control of this disease, because it is evident that not only humans but also animals should be surveyed to obtain an accurate understanding of parasite prevalence. As with xenomonitoring, animals can also be routinely surveilled to estimate parasite prevalence and the risk of LF becoming endemic in those areas.

## Conclusions

This study analyzed the morphological, morphometric parameters, and phylogenetic relationship of the brugian parasite in humans in Sri Lanka and the zoonotic potential of this parasite. Morphological and morphometric analysis placed the parasite within the range of *B. malayi.* However, sequence homology and phylogenetic analysis demonstrated that the parasite is distinct from FR3 reference strains of *B. malayi* and *B. pahangi*. Analysis of the *COXΙ* region revealed a closer relationship towards the brugian parasite reported previously from Sri Lanka and India. And the *HhaΙ* region revealed a closer relationship to the *B. malayi*-like parasite from India. This observation is supported by an increased genetic closeness to South Asian brugian strains than Thai strains in the genetic divergence analysis.

The presence of SNPs suggestive of canine origin and the detection of both human and animal DNA within the same mosquito, provides compelling support to the zoonotic potential of this parasite, highlighting its public health significance. These findings emphasize the need for one health approach, where animals are also routinely screened in addition to humans and mosquitoes. Regular screening of domestic animals, particularly dogs, alongside enhanced entomological surveillance and appropriate screening of travelers and migrants arriving from endemic regions is recommended to better understand transmission dynamics and to minimize the risk of ongoing spread and zoonotic transmission.

## Data Availability

The minimal data set is provided within the manuscript. Existing data were obtained from the National Center for Biotechnology Information via https://www.ncbi.nlm.nih.gov/, and accession numbers are provided where applicable. All relevant data from this study will be made available upon study completion.

## Acknowledgments

The authors gratefully acknowledge Dr. Michael Kimber for bearing the major financial burden of the project, Dr. Matthew Brewer for valuable suggestions for improvement of the quality of the manuscript and the Organization for Women in Science for the Developing World (OWSD), Trieste, Italy for funding the training at Iowa State University, USA, where the majority of the molecular work was carried out. They also extend their sincere appreciation to the staff of the Anti-filariasis Campaign, especially Mrs. Lakmini Liyanage, the staff of the Regional Director of Health Services branches at Madampe, Colombo, Kalutara, and Galle, the staff of the Department of Entomology at the Medical Research Institute, and Mr. Lahiru Herath for supporting the field study.

## References

1. World Health Organization (WHO). Lymphatic Filariasis. 2024. Accessed on 25 Jan 2026. https://www.who.int/news-room/fact-sheets/detail/lymphatic-filariasis.

2. World Health Organization (WHO). Global programme to eliminate preventing disability. 2013. Accessed on 25 Jan 2026. https://www.who.int/teams/control-of-neglected-tropical-diseases/lymphatic-filariasis/global-programme-to-eliminate-lymphatic-filariasis.

3. Asiedu SO, Kwarteng A, Kobla E, Amewu A, Kini P, Aglomasa BC, et al. Financial burden impact quality of life among lymphatic filariasis patients. BMC public health. 2021;1–10. doi: 10.1186/s12889-021-10170-8.

4. Eneanya OA, Garske T, Donnelly CA. The social, physical and economic impact of lymphedema and hydrocele : a matched cross-sectional study in rural Nigeria. BMC Infect Dis. 2019;1–16. doi: 10.1186/s12879-019-3959-6.

5. Ramaiah KD, Das PK, Michael E, Guyatt H. The economic burden of Lymphatic Filariasis in India. Parasitol Today, 2000;16(6):251–3.

6. Babu B V, Nayak AN, Dhal K, Acharya AS, Jangid PK, Mallick G. The economic loss due to treatment costs and work loss to individuals with chronic lymphatic filariasis in rural communities of Orissa, India, Acta Tropica, 2002;82:31–8.

7. Gyapong JO, Gyapong M, Evans DB, Aikins MK, Adjei S, Gyapong M, et al. The economic burden of lymphatic filariasis in northern Ghana. Ann. Trp. Med. Parasitol. 1996; 90: 39–48.

8. Mathew CG, Bettis AA, Chu BK, English M, Ottesen EA, Bradley MH, et al. The Health and Economic Burdens of Lymphatic Filariasis Prior to Mass Drug Administration Programs. Clin. Infect. Dis. 2020;70(12):2561–7. doi: 10.1093/cid/ciz671.

9. Ratnatunga PCA, Dandeniya C, Kandappa N, Balasuriya N, Manorangi EG. Lymphoedema of the limbs : an experience from a tertiary clinic in the central province of Sri Lanka. Sci Art. 2017;35(2):1–6. doi: 10.4038/sljs.v35i2.8381.

10. Lambrecht FL. Entomological aspects of filariasis control in Sri Lanka Coiombos. Bull World Health Organ. 1974;133–43.

11. Yahathugoda TC, Wickramasinghe D, Weerasooriya MV, Samarawickrema WA. Lymphoedema and its management in cases of lymphatic filariasis : the current situation in three suburbs of Matara, Sri Lanka, before the introduction of a morbidity-control programme. Ann. Trp. Med. Parasitol. 2005;99(5):501–10. doi: 10.1179/136485905X46450.

12. Wijesinghe RS, Wickremasinghe AR. Transactions of the Royal Society of Tropical Medicine and Hygiene Quality of life in filarial lymphoedema patients in Colombo, Sri Lanka. R. Soc. Trop. Med. Hyg. 2010;104:219–24. doi: 10.1016/j.trstmh.2009.08.005.2010;104:219–24.

13. World Health Organization (WHO). Expert Mission to Sri Lanka for Verification of Elimination of Lymphatic Filariasis. 2011;(June).

14. Cano J, Rebollo MP, Golding N, Pullan RL, Crellen T, Soler A, et al. The global distribution and transmission limits of lymphatic filariasis : past and present. Parasit Vectors. 2014;1–19.

15. Lucas GN. Filariasis-free Sri Lanka. Vol. 46, Sri Lanka. J. Child Health. 2017;46(3): 201–2.

16. Mallawarachchi CH, Nilmini Chandrasena TGA, Premaratna R, Mallawarachchi SMNSM, De Silva NR. Human infection with sub-periodic *Brugia* spp. in Gampaha District, Sri Lanka: A threat to filariasis elimination status? Parasit Vectors. 2018;11(1):18–23. doi: 10.1186/s13071-018-2649-3.

17. Mallawarachchi CH, Chandrasena TGAN, Withanage GP, Premarathna R, Mallawarachchi SMNSM, Gunawardane NY, et al. Molecular Characterization of a Reemergent *Brugia malayi* Parasite in Sri Lanka, Suggestive of a Novel Strain. Biomed Res Int. 2021;2021. doi: 10.1155/2021/9926101.

18. Kuruppu KAAS, Wickramasinghe S, Samarasekera SD, Wijegunawardana NDAD. S L J C R - 2 0 1 4 Detection of Human filarial parasite *Brugia malayi* in dogs in Sri Lanka. Vet Parasitol. 2020;2014.

19. Mallawarachchi CH, Chandrasena, Wichramasinghe S, Premaratna R, Gunawardane NYIS, Mallawarachchi NSMSM et al. A preliminary survey of filarial parasites in dogs and cats in Sri Lanka. PLoS One. 2018;13:1–11. doi: 10.1371/journal.pone.0206633.

20. Ambily VR, Pillai UN, Arun R, Pramod S, Jayakumar KM. Detection of human filarial parasite *Brugia malayi* in dogs by histochemical staining and molecular techniques. Vet Parasitol. 2011 Sep 27;181(2–4):210–4. doi: 10.1016/j.vetpar.2011.04.041.

21. Azaziah P, Abd M, Irwin PJ, Gatne M, Coleman GT, Mcinnes LM, et al. A survey of canine filarial diseases of veterinary and public health significance in India. Parasit Vectors. 2010;11–3. doi: 10.1186/1756-3305-3-30.

22. Gunaratna IE, Huggins LG, Atapattu U, Liyanage L, Shilpeswarage N, Vallipuranathan M, et al. Emergence of a novel zoonotic brugian filarial infection during post-validation surveillance for lymphatic filariasis in Sri Lanka. Lancet Reg Heal - Southeast Asia. 2026;46:100735. doi:10.1016/j.lansea.2026.100735.

23. Atapattu U, Koehler A V., Huggins LG, Wiethoelter A, Traub RJ, Colella V. Dogs are reservoir hosts of the zoonotic *Dirofilaria* sp. ‘hongkongensis’ and potentially of Brugia sp. Sri Lanka genotype in Sri Lanka. One Heal. 2023;17(August):100625. 10.1016/j.onehlt.2023.100625.

24. Nimalrathna SU, Harischandra H, Chandrasena N, Kimber MJ, Silva N De, Mallawarachchi CH, et al. Post elimination of lymphatic filariasis: a situation analysis of brugian filariasis and vector potentialities within the filarial transmission belt in Sri Lanka. Parasit Vectors. 2026;1–12.

25. Orihel TC, World Health Organization. Division of Control of Tropical Diseases. Bench aids for the diagnosis of filarial infections. Division of Control of Tropical Diseases, World Health Organization; 1997. 5.

26. Huat L, Yik M, Mahmud R, Muslim A. Lau YL, Kamarulzaman A, Zoonotic *Brugia pahangi* filariasis in a suburb of Kuala Lumpur City, Malaysia. Parasitol Int. 2011;60(1):111–3. doi: 10.1016/j.parint.2010.09.010.

27. Partono F. A Polymerase detection chain reaction assay for the of *Brugia malayi* in blood. Am. J. Trop. Med. Hyg. 1994;51(3):314–21. doi: 10.4269/ajtmh.1994.51.314.

28. Sanger F, Nicklen S, Coulson AR. DNA sequencing with chain-terminating inhibitors. Proc Natl Acad Sci U S A. 1977;74(12):5463–7.

29. Pitzer JB, Kaufman PE, Tenbroeck SH, Maruniak JE. Host blood meal identification by multiplex polymerase chain reaction for dispersal evidence of stable flies (Diptera: Muscidae) between livestock facilities. J Med Entomol. 2011;48(1):53–60.

30. Yen PKF, Mak JW. Histochemical differentiation of *Brugia*, *Wuchereria*, *Dirofilaria* and *Breinlia* microfilariae. Ann. Trop. Med. Parasitol. 2017;4983(April). doi: 10.1080/00034983.1978.11719298

31. Schacher JF. Morphology of the Microfilaria of *Brugia pahangi* and of the Larval Stages in the Mosquito. J. Parasitol. 1962;48(5):679–92.

32. Evans CC, Greenway KE, Campbell EJ, Dzimianski MT, Mansour A, Mccall JW, et al. The Domestic Dog as a Laboratory Host for *Brugia malayi*. Pathogens. 2022;11. doi: 10.3390/pathogens11101073

33. Laing ABG, Edeson JFB, Wharton RH. Studies on Filariasis in Malaya: The Vertebrate Hosts of *Brugia malayi* and *B.pahangi*. Ann Trop Med Parasitol, 2017;4983: 92–9. doi: 10.1080/00034983.1960.11685961.

34. Rajapakshe R, Perera WSR, Ihalamulla RL, Weerasena KH, Jayasinghe S, Sajeewani HBR. Study of dirofilariasis in a selected area in the Western Province. Ceylon Med. J., 2004:58–61.

35. Dissanaike AS, Jayaweera Bandara CD, Padmini HH, Ihalamulla RL, Naotunne TDS. Recovery of a species of *Brugia*, probably *B. ceylonensis*, from the conjunctiva of a patient in Sri Lanka. Ann Trop Med Parasitol. 2000;94(1):83–6. DOI: 10.1080/00034983.2000.11813516.

36. Buckley JJC, Nelson GS, Heisch RB. On *Wuchereria patei* n.sp. from the Lymphatics of Cats, Dogs and Genet Gats on Pate Island, Kenya. J. Helminthol. 1957;XXXII:73–80.

37. Ash ALR, Little MD. Linked references are available on JSTOR for this article : *Brugia beaveri* sp. n. (Nematoda : Filarioidea) from the Raccoon (Procyon lotor) in Louisiana. J Parasitol. 1964;50(1):119–23.

38. Eberhard ML, Telford, Andrew S. A *Brugia* species infecting rabbits in the northeastern United States. J Parasitol. 1991;77(5):796–8.

39. Orighel TC, *Brugia tupaiae* sp. n. (Nematoda: Filarioidea) in Tree Shrews (*Tupaia glis*) from Malaysia. J Prasitol. 2025;52(1):162–5.

40. Orihel TC. *Brugia guyanensis* sp. n. (Nematoda: Filarioidea) from the Coatimundi (*Nasua nasua viffata*) in British Guiana. J Parsitol. 2025;50(1):115–8.

41. Purnomo. Dennis DT, Partono F. The Microfilaria of *Brugia timori* (Partono et al. 1977 = Timor Microfilaria, David and Edeson, 1964): Morphologic Description with Comparison to *Brugia malayi* of Indonesia. J Parasitol, 2025;63(6):1001–6.

42. Dissanaike AS, Paramananthan DC. On *Brugia* (*Brugiella* subgen. nov.) buckleyi n. sp., from the Heart and Blood Vessels of the Ceylon Hare. J. Helminthol. 1961;XXXV(1):209–20.

43. Qing X, Kulkeaw K, Wongkamchai S, Tsui SKW. Mitochondrial Genome of *Brugia malayi* Microfilariae Isolated From a Clinical Sample. Front Ecol Evol. 2021 Feb 2;9. doi: 10.3389/fevo.2021.637805.

44. Turner HC, Bettis AA, Chu BK, McFarland DA, Hooper PJ, Mante SD, et al. Investment Success in Public Health: An Analysis of the Cost-Effectiveness and Cost-Benefit of the Global Programme to Eliminate Lymphatic Filariasis. Clin Infect Dis. 2017;64(6):728–35. doi: 10.1093/cid/ciw835.

45. Diekmann I, Supali T, Fischer K, Iskandar E. *Brugia malayi* and other filarial parasite species in animals in areas endemic for lymphatic filariasis in Belitung District, Indonesia. PLoS Negl Trop Dis, 2025; 19(1):1–13. doi: 10.1371/journal.pntd.0013593.

46. Kanjanopas K. Choochote W. Jitpakdi A. Suwannadabba S. Loymak S. Chungpivat S., et al. *Brugia malayi* in a naturally infected cat from Narathiwat province, Southern Thailand. Southeast Asian J Trop Med Public Health. 2001;32(3):585–7.

47. Chirayath D, Alex PC, Pillai UN, George S, Ajithkumar S, Panicker VP. Identification of *Brugia malayi* in dogs in Kerala, India. Trop Biomed. 2017;34(4):804–14.

48. Nuchprayoon S, Sangprakarn S, Junpee A, Nithiuthai S, Chungpivat S PY. Differentiation of *Brugia malayi* and *Brugia pahangi* by PCR-RFLP of ITS1 and ITS2. Southeast Asian J Trop Med Public Heal. 2003;34(2):67–73.

49. Chansiri K, Tejangkura T, Kwaosak P, Sarataphan N, Phantana S, Sukhumsirichart W. PCR-based method for identification of zoonotic *Brugia malayi* microfilariae in domestic cats. Mol Cell Probes. 2002;16(2):129–35. doi: 10.1006/mcpr.2001.0402.

50. Yilmaz E, Fritzenwanker M, Pantchev N, Lendner M, Wongkamchai S, Otranto D, et al. The Mitochondrial Genomes of the Zoonotic Canine Filarial Parasites *Dirofilaria (Nochtiella) repens* and *Candidatus Dirofilaria (Nochtiella)* Honkongensis provide evidence for presence of cryptic species. PLoS Negl Trop Dis. 2016;10(10):1–22.

51. Strübing U, Lucius R, Hoerauf A, Pfarr KM. Mitochondrial genes for heme-dependent respiratory chain complexes are up-regulated after depletion of *Wolbachia* from filarial nematodes. Int. J. Parasitol. 2010;40:1193–202.doi: 10.1016/j.ijpara.2010.03.004.

52. Duret L. Neutral Theory: The Null Hypothesis of Molecular Evolution. Nat Educ. 2008;1(1):218.

53. Saeed M, Siddiqui S, Bajpai P, K. Srivastava A, Mustafa H. Amplification of *Brugia malayi* DNA using Hha1 Primer as a Tool. Open Conf Proc J. 2015;5(1):38–40.

54. Tracey A, Foster JM, Paulini M, Grote A, Mattick J, Tsai Y chih, et al. Nearly Complete Genome Sequence of *Brugia malayi* Strain FR3. Microbiol Resour Announc. 2020;3–5.

55. Ravindran R, Varghese S, Nair SN, Balan VM, Lakshmanan B, Ashruf RM, et al. Canine filarial infections in a human *Brugia malayi* endemic area of India. Biomed Res Int. 2014;2014. doi: 10.1155/2014/630160.

56. Jayewardene LG. On Two Filarial Parasites from Dogs in Ceylon, Brugia ceylonensis n.sp. and Dipetalonema sp.inq. J Helminthol. 1962;36(3):269–80.

57. Anti Filariasis Campaign, Ministry of Health, Sri Lanka. Annual Health Bulletin 2022.

58. Anti Filariasis Campaign, Ministry of Health, Sri Lanka. Annual Health Bulletin 2023.

